# Development and validation of the long covid symptom and impact tools, a set of patient-reported instruments constructed from patients’ lived experience

**DOI:** 10.1101/2021.03.18.21253903

**Authors:** Viet-Thi Tran, Caroline Riveros, Bérangère Clepier, Moïse Desvarieux, Camille Collet, Youri Yordanov, Philippe Ravaud

**Author notes:** **Correspondence to:** Viet-Thi Tran - MD, PhD, Hôpital Hôtel Dieu, Centre d’Épidémiologie Clinique, Paris, France, 1 place du Parvis Notre-Dame, Paris 75181, France.

## Abstract

**Objectives:** To develop and validate patient-reported instruments, based on patients’ lived experiences, for monitoring the symptoms and impact of long covid.

**Design:** The long covid Symptom and Impact Tools (ST and IT) were constructed from the answers to a survey with open-ended questions to 492 patients with long covid. Validation of the tools involved adult patients with suspected or confirmed covid-19 and symptoms extending over three weeks after onset. Construct validity was assessed by examining the relations of the ST and IT scores with health related quality of life (EQ-5D-5L), function (PCFS, post-covid functional scale), and perceived health (MYMOP2). Reliability was determined by a test-retest. The “patient acceptable symptomatic state” (PASS) was determined by the percentile method.

**Results:** Validation involved 1022 participants (55% with confirmed covid-19, 79% female and 12.5% hospitalised for covid-19). The long covid ST and IT scores were strongly correlated with the EQ-5D-5L (r_s_ = −0.45 and r_s_ = −0.59 respectively), the PCFS (r_s_ = −0.39 and r_s_ = −0.55), and the MYMOP2 (r_s_ = −0.40 and r_s_ = −0.59). Reproducibility was excellent with an interclass correlation coefficient of 0.83 (95% confidence interval 0.80 to 0.86) for the ST score and 0.84 (0.80 to 0.87) for the IT score. In total, 793 (77.5%) patients reported an unacceptable symptomatic state, thereby setting the PASS for the long covid IT score at 30 (28 to 33).

**Conclusions:** The long covid ST and IT tools, constructed from patients’ lived experiences, provide the first validated and reliable instruments for monitoring the symptoms and impact of long covid.

**Short summary:** We developed the long covid Symptom and Impact Tools (ST and IT) from the experiences of 492 patients, captured during a survey with open-ended questions, and assessed their validity and reliability in a sample of 1022 patients with long covid.

## Introduction

As of February 2021, about 100 million people worldwide have been infected by the SARS-CoV 2 virus.^1^ According to some studies, from 10% to 70% of them, including many who only had a mild disease, report the presence of symptoms such as fatigue, dyspnoea, chest pain, memory problems, and concentration disorders persisting beyond three weeks from the initial onset of symptoms.^2-8^ Yet, so far, surveillance systems around the world remain focused exclusively on the acute aspect of the pandemic, tracking new cases, hospitalisations, and deaths, while neglecting these long-term consequences of covid-19.^9^

Persisting signs and symptoms three weeks after the acute covid-19 infection may be associated with several distinct clinical entities: 1) organ injury from the acute disease or its treatment (e.g., lung scarring, complications of intensive care, etc.),^8 10-12^ 2) post-viral fatigue syndrome,^13-15^ 3) prolonged viral shedding, especially in immunosuppressed patients,^16^ 4) reinfection, and 5) mental factors, such as post-traumatic stress. From the patient’s perspective, all or several of these causes are often intertwined and produce a substantial burden of disease. As a result, patients themselves created the term “long covid” to describe all of the symptoms they could experience, independently of their specific cause.^17^

Today, advances in understanding and treating long covid are impeded by the heterogeneity of its assessment: each study uses different sets of symptoms, most generally defined from the care givers’ perspective.^3 18 19^ There is currently no comprehensive, validated, and reliable instrument for monitoring either the course of long covid symptoms or its impact on patients’ lives—an absence that incurs the risk of missing important patient perspectives. For example, many patients with long covid report an inability to sleep, difficulties concentrating, or exhaustion after mild exercise—manifestations not considered in all existing studies.^20^ The National Institute for Health and Care Excellence (NICE) recently called for the development and validation of scales to assess long covid,^21^ and in February 2021, the World Health Organization issued a standardised, albeit non-validated, case report form for the follow-up of patients after acute covid-19.^22^

In this study, we aimed to develop and validate a set of patient reported instruments for monitoring the symptoms and impact of long covid, usable in clinical practice and constructed from patients’ lived experience.

## Methods

The development and validation of these instruments applied a two-stage method.^23^ The study was nested within ComPaRe (Communauté de Patients pour la Recherche, www.compare.aphp.fr), an e-cohort of patients with chronic conditions who volunteer to participate in research by regularly answering patient reported outcome measures (PROMs) and patient reported experience measures (PREMs).^24^ Because of the similarities between long covid and chronic conditions, recruitment in ComPaRe was extended to patients reporting symptoms of covid-19 lasting more than three weeks past initial onset. All participants provide electronic consent before participating. The Institutional Review Board of Hôtel-Dieu Hospital, Paris, approved ComPaRe (IRB: 0008367).

### Stage 1: Development of a set of patient reported instruments to monitor the symptoms and impact of long covid from the patients’ lived experiences

We sought to develop the contents of the instruments from the patients’ lived experiences of long covid. We invited adult patients in the ComPaRe cohort who reported a covid-19 infection (laboratory confirmed or not) with symptoms persisting more than three weeks past the initial infection to complete an online survey of open-ended questions that asked them to describe in detail the symptoms, potential triggers, and the effects of long covid on their lives. The survey, which took place from October 14 to November 29, 2020, used broad questions to avoid directing patients about long covid’s consequences (**Supplementary material 1**).

Participants were recruited through a social media and media campaign and by partner patient associations. Participants who had participated were invited to encourage people in their entourage who may have had a COVID-19 infection, to participate, by a ‘snowball’ sampling method ^25^.

Two investigators (VTT and CR) used an inductive multiple-round qualitative content analysis to examine the participants’ responses. First, they independently coded the responses (i.e., they identified within each response any expression found in the text expressing a symptom or impact of long covid and assigned it a code). Answers related to the acute phase of covid-19 (that is, symptoms during the first three weeks of infection) were removed from analysis. For example, when a participant stated that “I had a cough and fever for one week and joint pain for 7 months”, we considered only “joint pain” to be a potential long covid symptom. If the response did not specify a timeframe, all reported symptoms were kept in the analysis. Next, the investigators grouped the identified codes into a standardised set of symptoms and consequences of long covid, based on their medical knowledge and the literature.^4 18^ Analysis continued on until a comprehensive list of symptoms was obtained, i.e., until data saturation was reached. The point of data saturation was assessed with a mathematical model to predict the number of new symptoms that could be identified by adding new participants to the study.^26^ In a third step, they reduced the number of symptoms by: 1) grouping closely related symptoms (eg, loss of taste and changes in taste were grouped as loss/change of taste) and 2) eliminating those expressed by less than 2% of participants in the open-ended survey.

The two investigators used the results of the open-ended survey to develop a preliminary patient reported instrument for monitoring long covid’s current symptoms and impact. We chose to split the instrument into two independent parts. The first part aimed at assessing patients’ long covid-related symptoms over the previous 30 days with the checklist of symptoms identified above, a list we named the long covid Symptom Tool (ST). The second part aimed at assessing the disease’s impact on their lives over the past 30 days from the responses to six questions constructed from themes identified and examples provided by patients in their open-ended answers. Items used a numeric scale ranging from 0 (no impact) to 10 (maximal impact). This second instrument was called the long covid Impact Tool (IT). These two instruments were reappraised by the two author patients (BC and CC) for content validity, clarity, and wording during a telephone interview with the main investigator (VTT) that used the double interview method.^27^

### Stage 2: Validation of the long covid symptom and impact tools

A second sample of adult patients reporting a SARS-CoV 2 infection (laboratory confirmed or not) with symptoms persisting three weeks past the initial infection served as the validation sample. Patients who had participated in the first stage could also participate in the validation stage. In addition, we increased our sample through a call for participation on the “TousAntiCOVID” app, the official French contact tracing app used by 12 million people. As our instrument intends to evaluate patients’ current symptoms and their impact, only patients reporting at least one symptom during the previous 30 days could participate in the validation study.

Each tool was validated independently. The long covid ST score was defined as the number of symptoms reported by patients, and the long covid IT score as the sum of the responses to each impact question.

Construct validity was determined by confirming several theories or conceptions (ie, constructs) about long covid. First, we hypothesised that both the ST and IT scores would be negatively correlated with patients’ quality of life, which was evaluated with the EuroQol five-dimension five-level (EQ-5D-5L) questionnaire and the EuroQol visual analogue scale (EQ-VAS).^28^ Our second hypothesis was that both the ST and IT scores would be positively correlated with patients’ functional status after covid-19, assessed by the Post-Covid Functional Scale (PCFS), an ordinal tool categorising patients in 5 grades ranging from 0 “No functional limitation” to 4 “Severe functional limitations”.^29 30^ Finally, we hypothesised that both the ST and IT scores would also be negatively correlated with patients’ general perceived health, assessed with the Measure Yourself Medical Outcome Profile 2 (MYMOP2) tool, an instrument focused on complaints that patients identified as most important to them.^31^ Correlations were assessed by Spearman correlation coefficients (r_s_) and considered high with r_s_ > 0.50 and moderate with rs = 0.35 to 0.50.

The reliability of both the ST and IT scores was determined by the test-retest method. Some patients completed the questionnaires twice: at baseline and two weeks later. Reproducibility was assessed by the intraclass correlation coefficient (ICC) for agreement.^32^ The 95% confidence intervals (95% CIs) were determined by a bootstrap method. Reproducibility was considered acceptable with ICC > 0.70.^33^ Reliability was also tested by Bland and Altman plots, which present the differences between two measurements against the means of the two measurements.^34^ Finally, internal consistency was assessed by Cronbach’s α and was considered acceptable between 0.70 and 0.95.^35^

The Patient Acceptable Symptom State (PASS) is the level of a continuous treatment outcome measure below which patients consider themselves well.^36^ The PASS for the long covid IT was determined by matching the scores to an anchor question: “*Taking into account all your symptoms in daily life and your functional impairment, do you consider that your current state is satisfactory?*” The threshold was the impact score below which 75% of patients considered their symptom state acceptable.^37^ Percentile bootstrap with 2000 replications provided the 95% CIs.

Statistical analyses were performed with R v. 3.6.3.

### Patient and public involvement

The tools were developed from qualitative data about patients’ lived experiences, captured during a survey with open-ended questions. The qualitative results and the implementation of the items in the patient reported tools were discussed in detail with two patients meeting the criteria for long covid. They also participated in the critical revision of the manuscript and are co-authors of the paper (BC and CC).

## Results

### Step 1: Development of the long covid symptom and impact tools from patients’ lived experiences

During the survey period (from October 14 to November 29, 2020), 528 members of the ComPaRe cohort received emails inviting them to complete the open-ended survey, and 492 (93%) did so. Their median age was 45 years (IQR 35 to 50.25), and 414 (84%) were women. In total, 210 (43%) had tested positive for SARS-CoV2 by PCR swab or serological assay and were considered confirmed covid-19 patients. Median time since symptom onset was 217 days (IQR 205 to 230) **(Supplementary material 2**).

Participants’ open-ended answers represented a corpus of 116 657 words. Inductive content analysis of patients’ answers was stopped after the analysis of 380 answers and the definition of 53 symptoms of long covid because models showed that 99.99% of all possible symptoms had been identified in both the total sample and in the subgroups of confirmed and suspected cases (**Supplementary material 3**). The list of symptoms was further organised in 10 categories: general symptoms (n=11), thorax (n=6), digestive (n=3), ear/nose/throat (n=5), eyes (n=3), genitourinary (n=2), hair and skin (n=4), musculoskeletal (n=4), neurological (n=11), and blood and lymph circulation (n=4) symptoms (**Supplementary material 4**).

The analysis of patients’ open-ended answers also enabled us to identify six aspects of patients’ lives that were affected by long covid: 1) difficulties in performing personal activities such as driving, shopping, or doing household chores; 2) difficulties in their professional lives; 3) difficulties in fulfilling their family roles and/or feeling that they’re a burden to their family or friends; 4) difficulties related to social activities; 5) morale, fear of the future and of not returning to normal, and 6) negative impact on relationships with care providers. Examples of what patients wrote are provided in **Supplementary material 5**.

Thus the list of symptoms and the themes and examples related to the impact of long covid on patients’ lives were used to develop preliminary versions of the long covid tools, which were further reviewed and improved by two patients. The **Box** presents the items for both tools.

### Step 2: Validation of the long covid symptom and impact tools

From November 30, 2020, to February 15, 2021, 1360 ComPaRe participants reported they had had long covid. In all, 1022 had experienced at least one covid-19-related symptom in the 30 days before responding to the survey and were included in the validation sample. Among them, 418 (40%) had participated in the development sample, in the first stage. **Table 1** presents the characteristics of all patients included in the validation sample. Their median age was 45 years (IQR 37 to 52), and 817 (79.9%) were women. Overall, 564 (55%) had tested positive for SARS-CoV2 by PCR swab or serological assay and were considered confirmed cases. Among all cases, 128 (12.5%) patients had been hospitalised for covid-19, and 17 (1.7%) had been admitted to an ICU. The median time since symptom onset was 263 days (IQR 123 to 289), with a bimodal distribution corresponding to the first (260 to 300 days) and second waves (50 to 100 days) in France.

**Table 1:** Characteristics of patients included in the validation step (n=1022)

| Characteristic | Total<br>(n=1022) | Confirmed<br>covid-19<br>(n=564)* | Suspected<br>covid-19<br>(n=458) |
| --- | --- | --- | --- |
| Age, median (Q1–Q3) – yr | 45<br>(37–52) | 46<br>(37.75–53) | 43<br>(37–51) |
| Male sex – no (%) | 205 (20.1) | 123 (21.8) | 82 (17.9) |
| **Educational level – no (%)** |  |  |  |
| Middle school or equivalent | 58 (5.7) | 30 (5.3) | 28 (6.1) |
| High school or equivalent | 128 (12.5) | 61 (10.8) | 67 (14.6) |
| Associate's degree | 213 (20.8) | 126 (22.3) | 87 (19.0) |
| Higher education | 601 (58.8) | 336 (59.6) | 265 (57.9) |
| Other | 22 (2.2) | 11 (2.0) | 11 (2.4) |
| **Comorbidities – no (%)** |  |  |  |
| High blood pressure | 35 (3.4) | 23 (4.1) | 12 (2.6) |
| Diabetes | 18 (1.8) | 13 (2.3) | 5 (1.1) |
| Stroke or cardiac ischaemic disease | 3 (0.3) | 2 (0.4) | 1 (0.2) |
| Asthma / COPD | 81 (7.9) | 38 (6.7) | 43 (9.4) |
| Cancer | 8 (0.8) | 8 (1.4) | 0 (0) |
| Depression/Anxiety | 39 (3.8) | 22 (3.9) | 17 (3.7) |
| Fibromyalgia | 20 (2.0) | 6 (1.1) | 14 (3.1) |
| **Time since symptom onset, median (Q1–Q3) - days** | 263<br>(123-289) | 234<br>(82-280) | 270<br>(260-299) |
| **Hospitalised for covid-19 – no (%)** | 128 (12.5) | 76 (13.5) | 52 (11.4) |
| **Hospitalised in ICU for covid-19 – no (%)** | 17 (1.7) | 13 (2.3) | 4 (0.9) |
| **Duration of hospitalisation, median (Q1-Q3)** | 4 (1–7.25) | 5 (2–9.25) | 2.5 (1–5.25) |

\*Reported positive testing for SARS-CoV2 by PCR swab or serological assay.

#### Symptoms and impact of long covid

The median long covid ST score was 16 (IQR 11 to 23). This score reports the number of symptoms patients experienced over the last 30 days and has a theoretical range from 0 (no symptoms) to 53 (all symptoms identified during step 1). The symptoms most frequently reported were fatigue (n=899), headaches (n=709), difficulties concentrating/mental fog (n=650), sleep disorders (n=603), and dyspnoea (n=570) (**Figure 1)**. Symptom frequency was similar in confirmed and suspected cases except for “change/loss of taste” and “change/loss of smell”, both more frequent in confirmed cases. Besides symptoms, 824 patients (80.6%) reported a relapsing-remitting disease course with daily (n=285, 34.6%), weekly (n=320, 38.8%), or less than weekly (n=219, 26.6%) relapses.

**Figure 1:**
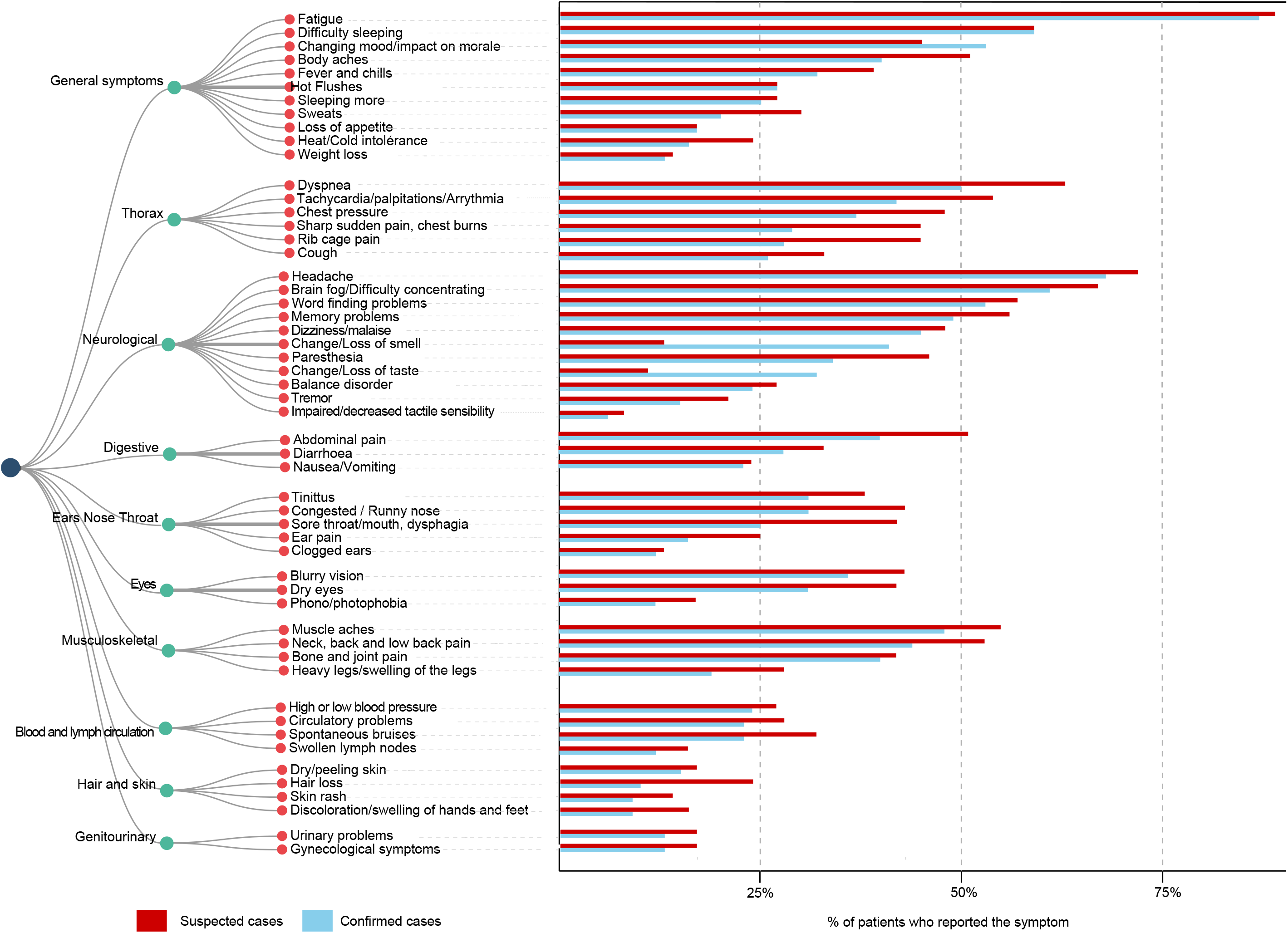
Symptoms reported by patients during the validation step (n=1022)

Patients’ median long covid IT score was 36 (IQR 24 to 45). This score has a theoretical range of 0 (no impact) to 60 (maximum impact) and represents the sum of item scores for the six questions related to the disease’s impact on their personal activities, family lives, professional lives, social lives, their morale, and their relationships with care providers. In all, 265 (26%) patients rated the impact of the disease on their work lives at 10 (out of 10). In general, the disease’s impact on patients’ lives was similar for patients with confirmed and suspected infections, except for the item about its effect on their relationships with care givers, which was rated higher in suspected cases (**Supplementary material 6**).

The long covid ST and IT scores were highly correlated (r_s_=0.54, p<0.0001) and did not seem to differ by time from symptom onset (**Supplementary material 7**).

#### Construct validity

Overall, 970 (95%) patients completed the EQ-5D-5L quality of life questionnaire, the EQ-VAS scale, and the PCFS functional status scale, while 746 (73%) answered the MYMOP2 questionnaire.

For health-related quality of life, the median EQ-5D-5L and EQ-VAS values in our sample were respectively 0.83 (IQR 0.60 to 0.89) and 51 (IQR 40 to 70). As hypothesised, we found that the long covid ST score was moderately and negatively correlated with the EQ-5D-5L questionnaire (r_s_ = −0.45, p<0.0001) and the EQ-VAS (r_s_ = −0.39, p<0.0001), while the long covid IT score had a strongly negative correlation with the EQ-5D-5L questionnaire (r_s_ = − 0.59, p<0.0001) and the EQ-VAS (r_s_ = −0.54, p<0.0001).

For functional assessments, 469 (48%) patients in our sample indicated that they were no longer able to perform some activities at home or at work by themselves (grade 3 or 4 of the PCFS), and 371 (38%) reported that they had had to reduce some of their activities (grade 2 of the PCFS). We found a moderate correlation between the long covid ST score and the PCFS score (r_s_ = −0.39, p<0.0001) and a high correlation between the long covid IT score and the PCFS score (r_s_ = −0.55, p<0.0001).

Similarly, for patients’ perceived health state, we found that the long covid ST score was moderately correlated with the MYMOP2 score (r_s_ = −0.40, p<0.0001) while the long covid IT score was highly correlated with it (r_s_ = −0.59, p<0.0001) (**Table 2**).

**Table 2:** Relations between patients’ long covid symptom tool (ST) score, impact tool (IT) score, quality of life, functional status, and perceived health (n=970)

| Characteristic | Relationship with the long covid ST score | p | Relationship with long covid IT score | p |
| --- | --- | --- | --- | --- |
| **Quality of life (EQ5D-5L)\*** | $r_s = -0.46$ | $<0.0001$ | $r_s = -0.59$ | $<0.0001$ |
| **Quality of life (EQ-VAS)\*** | $r_s = -0.39$ | $<0.0001$ | $r_s = -0.54$ | $<0.0001$ |
| **Post-covid functional status (PCFS)\*** | $r_s = 0.39$ | $<0.0001$ | $r_s = 0.55$ | $<0.0001$ |
| **MYMOP2 score\*** | $r_s = 0.40$ | $<0.0001$ | $r_s = 0.59$ | $<0.0001$ |

Relations between the scores and continuous variables were explored with Spearman's correlation coefficient. Correlation coefficients range from -1 (perfect negative correlation) to 1 (perfect positive correlation) with 0 indicating no correlation. \*Higher EQ5D scores indicate better quality of life; higher PCFS scores indicate more severe functional limitations; higher MYMOP2 scores indicate worse general health.

#### Reliability

Of the 351 patients invited to complete the long covid ST and IT twice for the test-retest, 235 (67%) did so. The symptom score had an ICC of 0.83 (95% CI 0.80 to 0.86), with Bland and Altman plots showed a mean difference of 0.8 (95% limits of agreement, −14 and 16). The impact score’s ICC was 0.84 (95% CI 0.80 to 0.87), with Bland and Altman plots showing a mean difference of 0.5 (95% limits of agreement, −11 to 12 (**Figure 2**). Finally, Cronbach’s alpha was 0.89 (95% CI, 0.88 to 0.90) for the long covid ST score and 0.86 (95% CI, 0.85 to 0.88) for the IT score.

**Figure 2:**
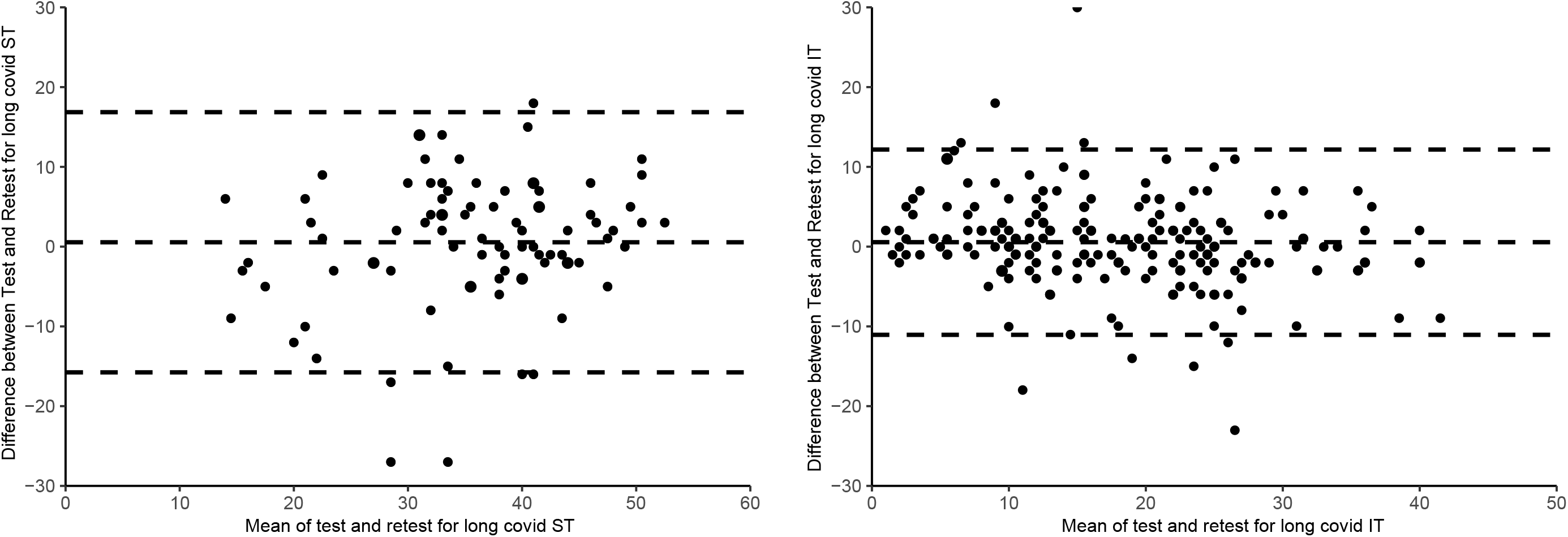
Bland and Altman plots for the test-retest of the long covid Symptom Tool score (A) and Impact Tool Score (B) scores (n=235)

#### Patient acceptable symptom state

Only 229 (22.4%) patients reported an acceptable symptomatic state, while 793 (77.5%) patients reported it was unacceptable. Long covid’s impact was considered acceptable for >75% of patients with IT scores < 30 (95% CI, 28 to 33), that is, below 50% of the maximum score. (**Figure 3**).

**Figure 3:**
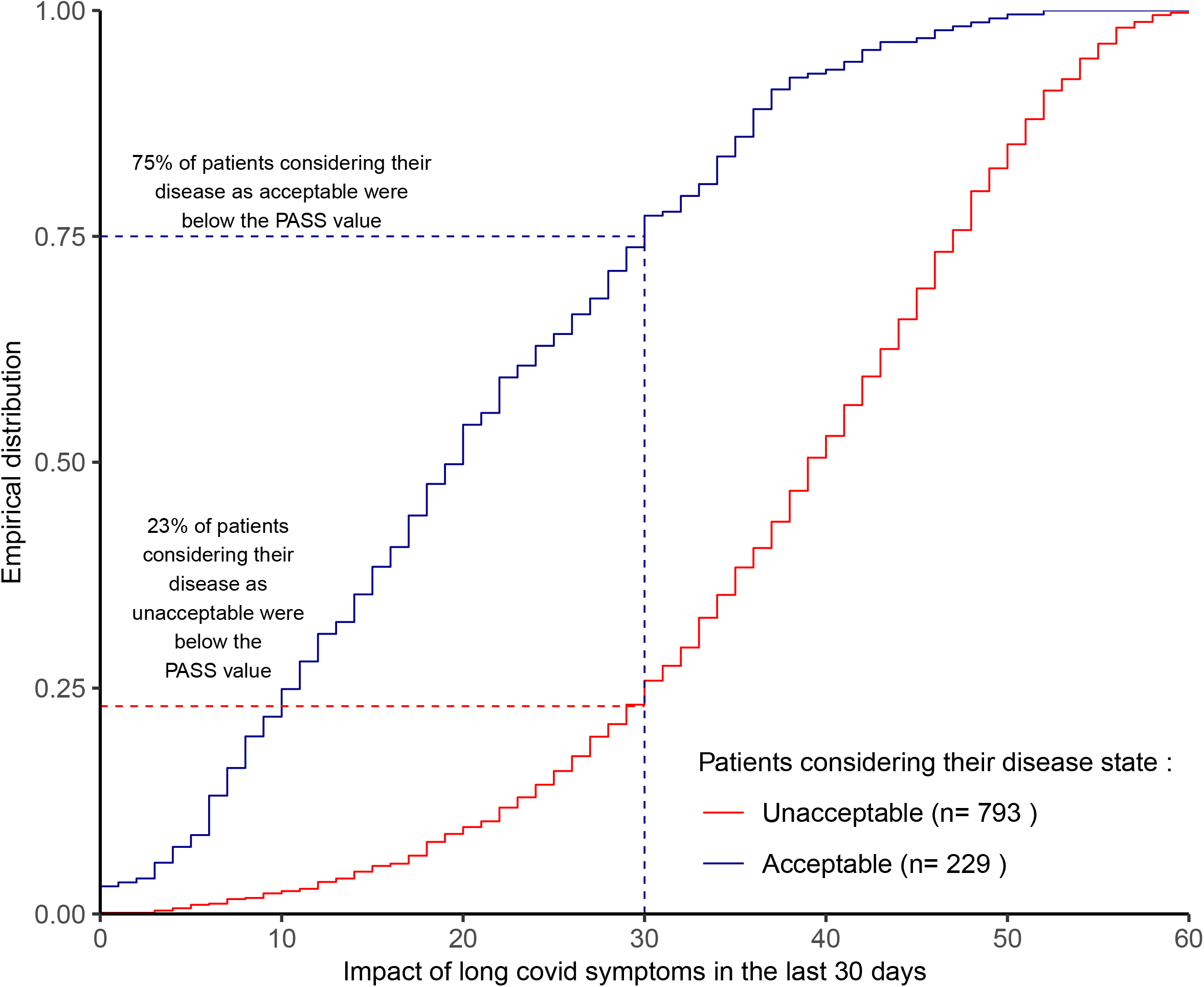
Empirical cumulative distribution function for long covid Impact Tool score, in patients considering their levels of symptoms and impact acceptable (n=229) and not acceptable (n=793)

## Discussion

In this study, we present the long covid ST and IT. They are the first set of validated and reliable instruments for monitoring this disease’s symptoms and impact and assess respectively 53 long covid symptoms and 6 dimensions of patients’ lives that it can affect.

Until now, two issues have limited the assessment of long covid. First, the sets of symptoms used have been both unreliable and defined from the care givers’ perspective. The number of symptoms assessed in studies has ranged from 8 to 205 manifestations, the latter also including triggers and laboratory test result anomalies^3 7 8 19 38^ and usually failed to cover difficulties in concentration, even though several studies report that this symptom is frequent in long covid.^8 38^ The long covid ST and IT were built from patients’ lived experiences, captured during a survey with open-ended questions. Our tools thus provide a comprehensive picture of the disease. By using mathematical models to assess data saturation, we ensured that our instruments covered all manifestations and impacts of long covid relevant to patients. Both of our tools showed robust psychometric properties. Construct validity was demonstrated by the high correlations with patients’ quality of life, functional status, and perceived health state. Reliability was excellent with an ICC ≥ 0.8 during the test-retest. Finally, we defined a PASS (Patient Acceptable Symptomatic State) for long covid that provides thresholds to be met by pharmacological and/or non-pharmacological interventions aimed at reducing its impact on patients’ lives.

Second, most studies have focused on counting symptoms, but have failed to assess the disease’s impact on patients’ lives.^4 5 7 18 19^ The long covid IT fills this gap by providing validated, and reliable questions for assessing its burden. This is critical, in view of the major impact that long covid has on the quality of life and functioning of the patients in our ComPaRe cohort. The health related quality of life for patients with long covid, assessed with the EQ-VAS, was on average 40% lower than in the reference general population,^39^ and similar to that of patients living with epilepsy or multiple sclerosis.^40^ Half reported impaired functioning, that is, they were no longer able to perform some activities unassisted at home or at work (PCFS grade 3 or 4), a figure similar to that described in the study by Davis et al.^38^

The symptoms captured in the long covid ST overlap with those presented in sections 2.5 and 2.6 of the recent case report form issued by the WHO for the follow-up of patients after their acute illness. The WHO form covers 45 symptoms of long Covid (all included in our instrument) and includes questions on patients’ functioning (e.g., washing themselves, dressing, ability to join in community activities, etc.).^22^ Our set of instruments goes further by inquiring about patients’ perceptions of the disease’s impact on their lives, roles, and relationships, beyond assessing whether they can or cannot perform specific activities. Moreover, besides collecting information, our instruments provide a scoring method producing valid and reproducible measurements of the disease- and patient-relevant cut-offs to interpret these measures. In all, the overlap of the long covid ST and IT with the WHO case report form strengthens the content validity of our instruments and provides a glimpse of the potential validity and reliability of the WHO form.

This study has limitations. Generalisation of the estimates obtained in this study must be cautious. Our study recruited volunteer patients who reported persistent symptoms. This might have selected younger, more educated, and more often female patients willing to share their experiences with others and/or with more severe conditions. Nonetheless, we were able to involve and validate our instrument in a diverse sample of participants by using a broad media campaign and a call for participation on the “TousAntiCOVID” app.^41^ Second, our study may not be appropriate for examining the longitudinal impact of the disease. Although we recruited patients with various times from symptom onset, the assessment of symptoms and impact was cross-sectional. Future work following up patients with standardised, validated tools is required to investigate the duration of the disease and the course of the symptoms over time. Finally, in view of the limited number of patients who were hospitalised in ICUs in our study, we cannot confirm that our tool is suitable for measuring the consequences of ICU on patients.

According to the United Kingdom Office for National Statistics, 10% of people who were infected with SARS-CoV-2 still experience symptoms after three months, including those whose acute infection was asymptomatic.^42^ With about 100 million cases of COVID-19 worldwide, long covid may well affect millions of patients. The severe burden of illness and the impairment of quality of life associated with long covid call for urgent research to understand this disease and to develop interventions to help patients. Using scientific, valid, and reliable measurements in these initiatives will enable the comparison and combination of study results.

## Conclusions

The long covid Symptom and Impact Tools, constructed from patients’ lived experience, provide the first validated, reliable instruments for monitoring the symptoms and impact of long covid. It may help the development of treatment strategies to mitigate the considerable burden of this disease.

## Data Availability

All data collected for the study, including individual participant data and a data dictionary is available for research purposes, under the rules of the ComPaRe e-cohort (https://compare.aphp.fr/)

## Acknowledgements

The authors thank Elise Diard and Isabelle Pane for their help in the survey development, Isabelle Pane for data management and Elise Diard for drafting the figures.

## Contributions

Generated the idea: VTT and PR, Conceived and designed the experiments: VTT, and PR, Collected data: VTT, YY and PR. Analysed data: VTT, CR, BC, CC; Wrote the first draft of the manuscript: VTT, Contributed to the writing of the manuscript: VTT, CR, BC, CC, YY and PR; ICMJE criteria for authorship read and met: VTT, CR, BC, CC, YY and PR. Agree with manuscript results and conclusions: VTT, CR, BC, CC, YY and PR. VTT is the guarantor, had full access to the data in the study, and takes responsibility for the integrity of the data and the accuracy of the data analysis.

## Transparency statement

VTT affirms that the manuscript is an honest, accurate, and transparent account of the study being reported; that no important aspects of the study have been omitted. There were no discrepancies from the study as originally planned.

## Dissemination to participants and related patient and public communities

The results of your study will be sent to research participants and partner patients’ associations, accordingly to the protocol of ComPaRe. In addition, we plan a press release for the study.

## Declaration of interests

The authors declare no competing interests and no financial associations that may be relevant or seen as relevant to the submitted manuscript.

The authors have no association with commercial entities that could be viewed as having an interest in the general area of the submitted manuscript.

## List of Supplemental files

- **Supplementary material 1: Open-ended questions used for the development of the tool (step 1)**
- **Supplementary material 2: Characteristics of patients included in the survey with open-ended questions (step 1) (n=492)**
- **Supplementary material 3: Data saturation for symptoms identified during the survey with open-ended questions**
- **Supplementary material 4: Symptoms identified in the survey with open-ended questions (n=380)**
- **Supplementary material 5: Examples of what patients wrote about the impact of long Covid on their lives (n=380)**
- **Supplementary material 6: Patients’ scores (median [interquartile range]) to the items assessing the impact of long Covid (n=1022)**
- **Supplementary material 8: Long covid Symptom Tool (ST) score, Impact Tool (IT) score and EQ-5D-5L scores over time (n=970) (n=970)**

**Box:**
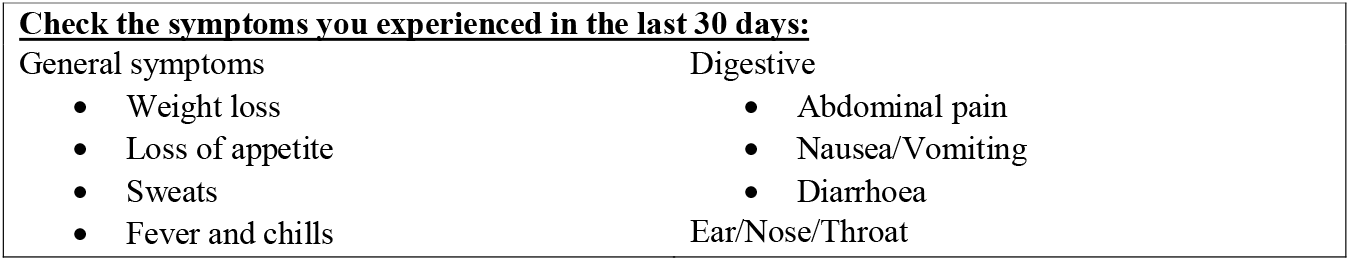
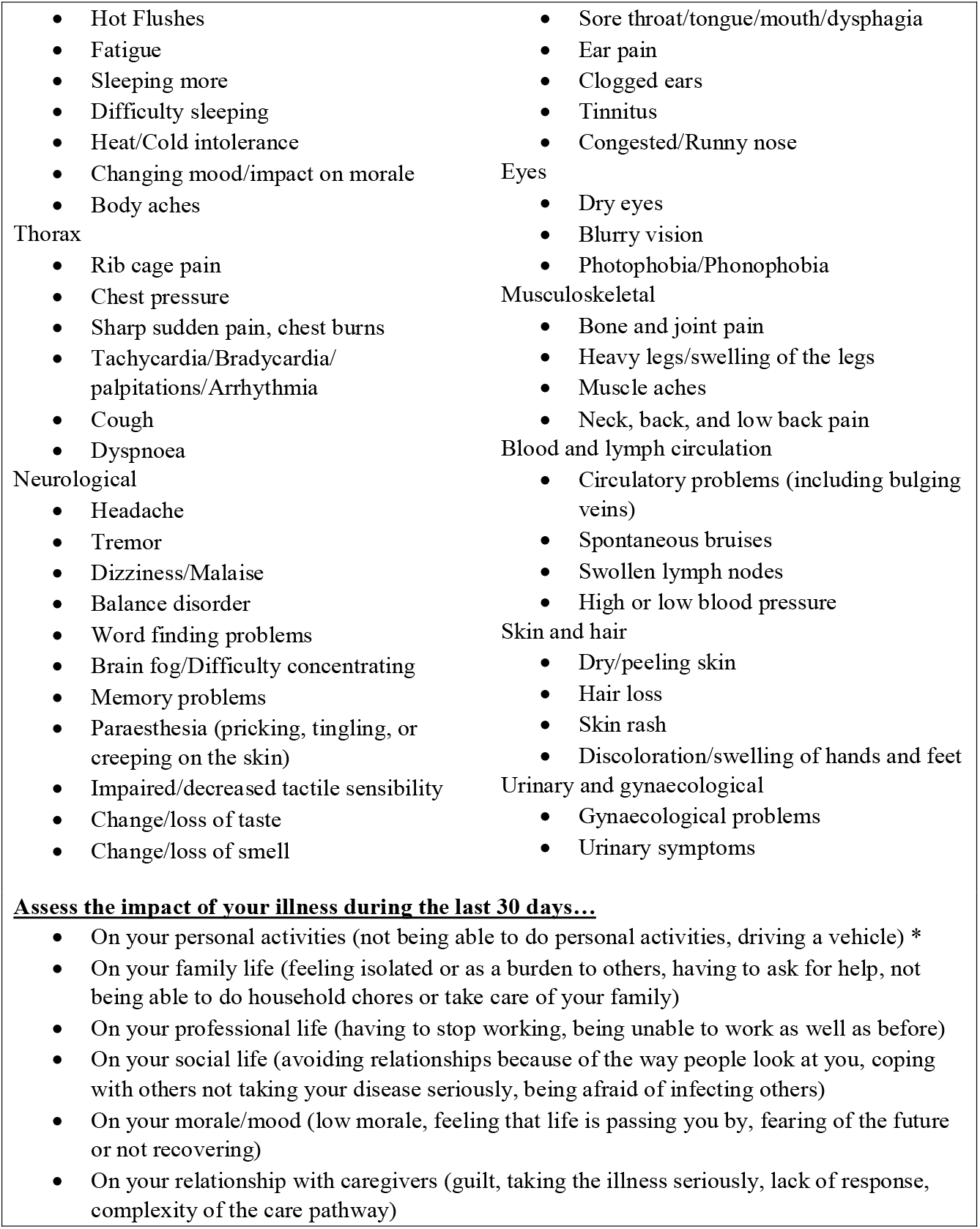
Items of the long covid symptom and impact tools (ST and IT)

