## Supplementary material for "Development and validation of the long covid symptom and impact tools, a set of patient-reported instruments constructed from patients’ lived experience"

### Supplementary material 1: Open-ended questions used for the development of the tool (step 1)

- Describe all the long-term symptoms of COVID-19 you have experienced. Do not hesitate to describe in detail these symptoms, including possible triggers and rhythm (relapse, etc.)
- Describe how long Covid is affecting your life in general.
- Describe how long Covid is affecting your professional life.
- Describe how long Covid is affecting your family and social life.
- Describe how long Covid is affecting your morale/mood.

### Supplementary material 2: Characteristics of patients included in the development of the tool (n=492)

| **Characteristic** | **Total**  **(n=492)** |
| --- | --- |
| **Age, median (Q1**−**Q3) – year** | 45  (35−50.25) |
| **Female sex – no (%)** | 414 (84) |
| **Educational level – no (%)**  Middle school or equivalent  High school or equivalent  Associate’s degree  Higher education  Other | 35 (7.1)  80 (16.3)  110 (22.4)  262 (53.3)  5 (1.0) |
| **Positive testing for SARS-CoV by PCR swab or serological assay – no (%)** | 210 (43%) |
| **Time since symptom onset, median (Q1**−**Q3) - days** | 217  (205-230) |
| **Hospitalized for COVID-19 – no (%)** | 56 (11.4) |
| **Hospitalized in ICU for COVID-19 – no (%)** | 3 (0.6) |
| **Duration of hospitalization, median (Q1-Q3)** | 3 (2-6) |

### Supplementary material 3: Data saturation for symptoms identified during the development of the tool (step 1). Plain line represents the cumulative curve of symptom identification during the qualitative step. The dotted line represents the potential symptoms that could be identified by analysing data from additional participants.

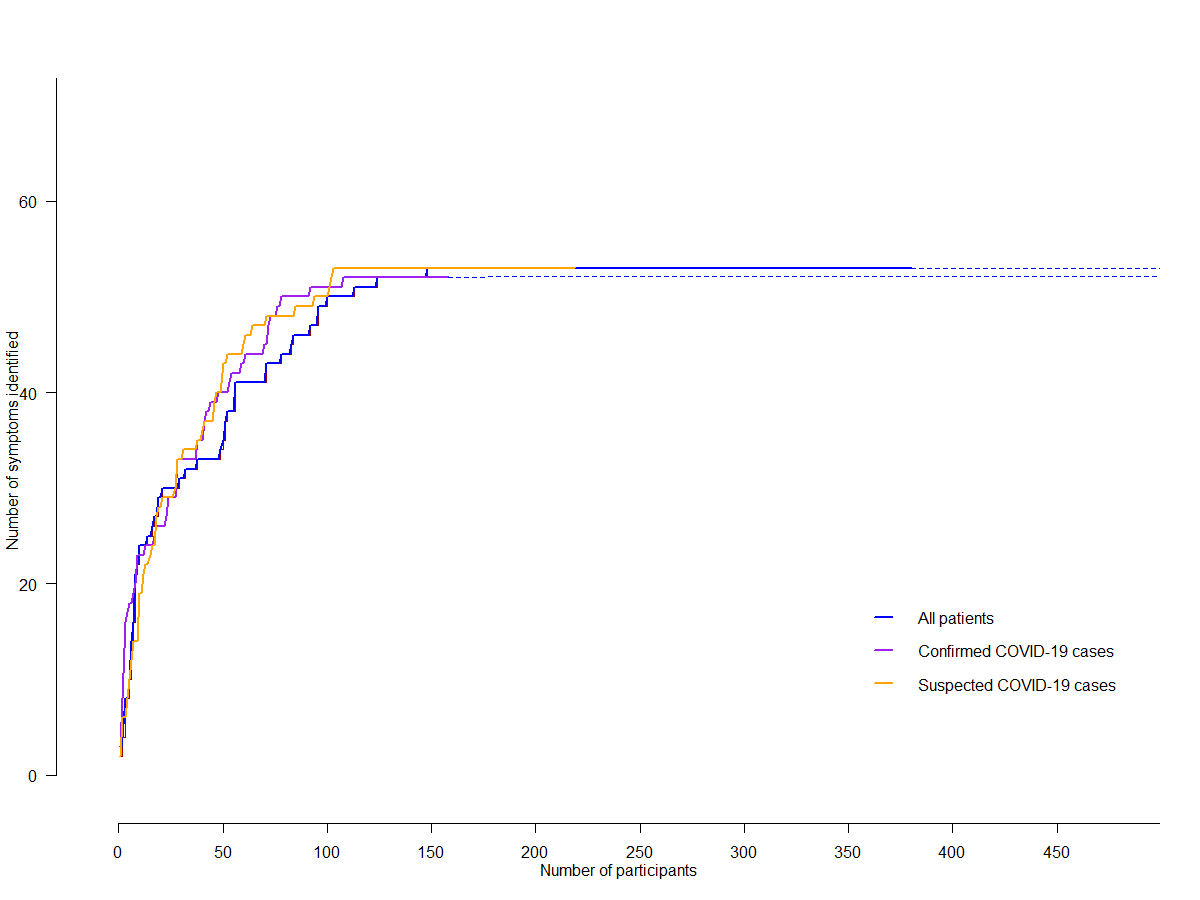

### Supplementary material 4: Symptoms identified in the survey with open-ended questions (n=380)

| **Symptoms identified in the survey with open-ended questions** | **Final list of symptoms retained** | **Category** |
| --- | --- | --- |
| Thirst/Hunger | *Eliminated because less than 2% of participants reported the symptom* | General symptoms |
| Weight loss | Weight loss |  |
| Weight gain | *Eliminated because less than 2% of participants reported the symptom* |  |
| Loss of appetite | Loss of appetite |  |
| Sweats | Sweats |  |
| Fever | Fever and chills |  |
| Chills (without fever) |  |  |
| Hot flushes | Hot flushes |  |
| Fatigue | Fatigue |  |
| Sleeping more | Sleeping more |  |
| Difficulty sleeping | Difficulty sleeping |  |
| Heat/Cold intolerance | Heat/Cold intolerance |  |
| Changing mood/impact on morale | Changing mood/impact on morale |  |
| Body aches | Body aches |  |
| Rib cage pain | Rib cage pain | Thorax |
| Chest pressure | Chest pressure |  |
| Sharp sudden pain, heartburn | Sharp sudden pain, heartburn |  |
| Tachycardia | Tachycardia/Bradycardia / palpitations/Arrhythmia |  |
| Bradycardia |  |  |
| Palpitations / Arrhythmia |  |  |
| Cough | Cough |  |
| Dyspnoea | Dyspnoea |  |
| Abdominal pain | Abdominal pain | Digestive |
| Nausea/Vomiting | Nausea/Vomiting |  |
| Diarrhoea | Diarrhoea |  |
| Constipation | *Eliminated because less than 2% of participants reported the symptom* |  |
| Sore throat/tongue/mouth | Sore throat/tongue/mouth/dysphagia | Ear/Nose/Throat |
| Dysphagia |  |  |
| Ear pain | Ear pain |  |
| Clogged ears | Clogged ears |  |
| Tinnitus | Tinnitus |  |
| Congested nose | Congested/runny nose |  |
| Runny nose |  |  |
| Dry eyes | Dry eyes | Eyes |
| Blurry vision | Blurry vision / |  |
| Floaters, light flashes | *Eliminated because less than 2% of participants reported the symptom* |  |
| Photophobia | Photophobia |  |
| Gynaecological problems | Gynaecological problems | Genitourinary |
| Urinary symptoms | Urinary symptoms |  |
| Dry/peeling skin | Dry/peeling skin | Hair and skin |
| Hair loss | Hair loss |  |
| Papulosquamous eruptions | Skin rash |  |
| Morbilliform |  |  |
| Discoloration/swelling of hands and feet | Discoloration/swelling of hands and feet |  |
| Bone pain | Bone and joint pain | Musculoskeletal |
| Joint pain |  |  |
| Heavy legs/swelling of the legs | Heavy legs/swelling of the legs |  |
| Muscle aches | Muscle aches |  |
| Neck, back and low back pain | Neck, back, and low back pain |  |
| Headache | Headache | Neurological |
| Tremor | Tremor |  |
| Convulsions | *Eliminated because less than 2% of participants reported the symptom* |  |
| Dizziness/Malaise | Dizziness/Malaise |  |
| Balance disorder | Balance disorder |  |
| Difficulty finding the right word | Word finding problems |  |
| Brain fog/Difficulty concentrating | Brain fog/Difficulty concentrating |  |
| Memory problems | Memory problems |  |
| Hallucinations | *Eliminated because less than 2% of participants reported the symptom* |  |
| Paraesthesia - pricking, tingling, or creeping on the skin | Paraesthesia — pricking, tingling, or creeping on the skin |  |
| Facial paralysis | *Eliminated because less than 2% of participants reported the symptom* |  |
| Impaired or decreased tactile sensibility | Impaired or decreased tactile sensibility |  |
| Change of taste | Change/loss of taste |  |
| Loss of taste |  |  |
| Change of smell | Change/loss of smell |  |
| Loss of smell |  |  |
| Circulatory problems (including bulging veins) | Circulatory problems (including bulging veins) | Blood and vessels |
| Spontaneous bruises | Spontaneous bruises |  |
| Swollen lymph nodes | Swollen lymph nodes |  |
| High or low blood pressure | High or low blood pressure |  |
| Anaemia | *Eliminated because is a laboratory value* |  |

### Supplementary material 5: Examples of what patients wrote about the impact of long covid on their lives (n=380)

| Theme identified | Example |
| --- | --- |
| Difficulties in performing basic activities | *I had to go back to live with my parents at the age of 31 because I am unable to carry out daily tasks”* (32-year-old woman) |
| Difficulties in their professional lives | “*I used to have a great capacity for work, but now I can no longer work full time [because of my symptoms] (…). This has an important economic impact on my life*” (55-year-old woman) |
| Difficulties in fulfilling their family roles | “*My family life is complex due to my permanent fatigue. I no longer have sex; sometimes my libido is totally absent.*” (40-year old man) |
| Difficulties in having social activities | “*I can't spend evenings with friends either because of the fatigue and the memory problems. I can no longer have long conversations*” (39-year-old woman) |
| Fear of missing out their lives and/or uncertainty about the future | “The *uncertainty about the duration of the illness is painful. Will I be able to stand up tomorrow? I can't tell colleagues if I'll be back in a week or a year. How to organize myself?*” (52-year-old woman) |
| Negative impact on relationships with care providers | “*My physician does not understand my problems and thinks I’m exaggerating my symptoms*” (60-year-old woman) |

### Supplementary material 6: Patients’ scores (median [interquartile range]) to the items assessing the impact of long Covid (n=1022)

| **Items** | **Total sample**  **(n=1022)** | **Confirmed cases**  **(n=564)** | **Suspected cases**  **(n=458)** |
| --- | --- | --- | --- |
| Difficulties in performing basic activities | 6 (3−7.5) | 6 (3−7) | 6 (4−8) |
| Difficulties in their professional lives | 7 (4−10) | 7 (3−10) | 7 (4−10) |
| Difficulties in fulfilling their family roles | 5 (3−7.5) | 5 (2−7) | 6 (3−8) |
| Difficulties in having social activities | 6 (3−8) | 5 (2−8) | 6 (4−8) |
| Fear of missing out their lives and/or uncertainty about the future | 7 (5−8) | 7 (5−8) | 7 (4−8) |
| Negative impact on relationships with care providers | 6 (3−8) | 5 (1−8) | 7 (4−9) |

*scores range from 0 (no impact) to 10 (maximal impact)

### Supplementary material 8: Long Covid Symptom Tool (ST) score, Impact Tool (IT) score and EQ-5D-5L scores over time (n=970)

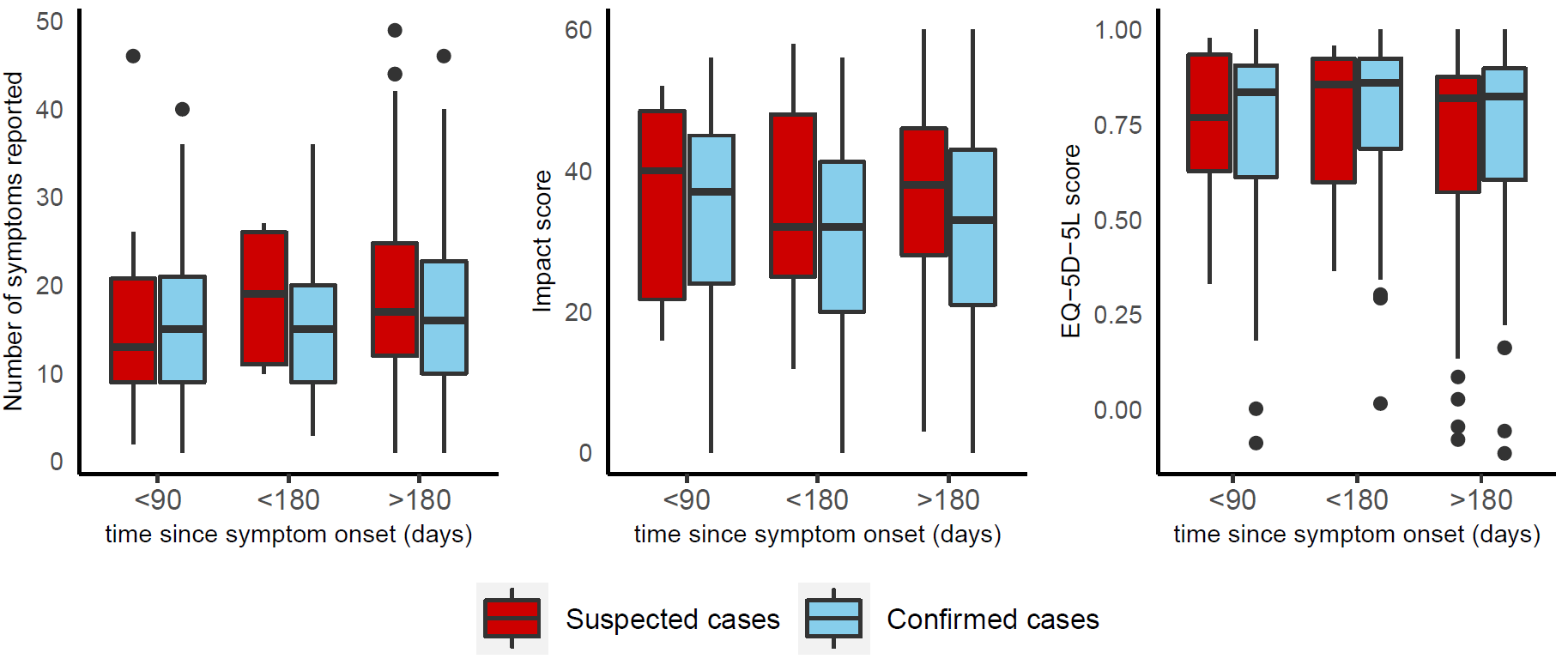
